# A novel comprehensive metric to assess COVID-19 testing outcomes: Effects of geography, government, and policy response

**DOI:** 10.1101/2020.06.17.20133389

**Authors:** Anthony C. Kuster, Hans J. Overgaard

**Author notes:** Corresponding author. (HJO).

## Abstract

Testing and case identification are key strategies in controlling the COVID-19 pandemic. Contact tracing and isolation are only possible if cases have been identified. The effectiveness of testing must be tracked, but a single comprehensive metric is not available to assess testing effectiveness, and no timely estimates of case detection rate are available globally, making inter-country comparisons difficult. The purpose of this paper was to propose a single, comprehensive metric, called the COVID-19 Testing Index (CovTI) scaled from 0 to 100, that incorporated several testing metrics. The index was based on case-fatality rate, test positivity rate, active cases, and an estimate of the detection rate. It used parsimonious modeling to estimate the true total number of COVID-19 cases based on deaths, testing, health system capacity, and government transparency. Publicly reported data from 188 countries and territories were included in the index. Estimates of detection rates aligned with previous estimates in literature (R^2^=0.97). As of June 3, 2020, the states with the highest CovTI included Iceland, Australia, New Zealand, Hong Kong, and Thailand, and some island nations. Globally, CovTI increased from April 20 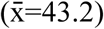 to June 3 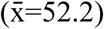 but declined in ca. 10% of countries. Bivariate analyses showed the average in countries with open public testing policies (59.7, 95% CI 55.6-63.8) were significantly higher than countries with no testing policy (30.2, 95% CI 18.1-42.3) (p<0.0001). A multiple linear regression model assessed the association of independent grouping variables with CovTI. Open public testing and extensive contact tracing were shown to significantly increase CovTI, after adjusting for extrinsic factors, including geographic isolation and centralized forms of government. This tool may be useful for policymakers to assess testing effectiveness, inform decisions, and identify model countries. It may also serve as a tool for researchers in analyses by combining it with other databases.

## Introduction

Coronavirus disease-2019 (COVID-19) is caused by infection of the Severe Acute Respiratory Syndrome Coronavirus 2 (SARS-CoV-2). The COVID-19 pandemic has forced many countries, states, and territories to enact public health measures to reduce its spread, including social distancing, contact tracing, stay-at-home orders, shuttering of schools, closure of public spaces, and border closures [1,2].

Testing, case identification, and isolation are critical activities to breaking the transmission chain [3]. Other measures, including social distancing and use of face masks, are also needed [4]. In order to assess testing and inform decisions about resuming economic activities, many countries and institutions have tracked testing-related metrics.

Thus far, the approach of many countries has generally been to specify several separate metrics related to testing, such as incidence, test positivity rate, number of hospitalizations, and mortality rate, with benchmarks for each criteria that are used to justify removing measures in phases [5,6]. However, many of these metrics rely on cases that have been identified through active diagnostic testing. One challenge, though, is the substantial proportion of the infected population that is asymptomatic [7,8]. Consequently, diagnostic testing has been inadequate to reveal what proportion of the population is infected, with real infections in most countries estimated to be 10 to 15 times, and sometimes even >100 times, higher than the reported number of cases [9–11].

Furthermore, predicating reopening dates on incidence may disincentivize testing, since increased diagnostic testing will inherently uncover more cases and, thus, delay reopening. Another criticism is that some criteria, such as the “downward trajectory,” specified by the US Centers for Disease Control [5], are vague. Thus, metrics used by policymakers and politicians to inform decisions should not only be quantitative but also encourage widespread proactive testing, such as the proportion of the total number of infections that have been detected [12]. However, estimating the detection rate/underreporting is challenging.

The level of undetected cases has been estimated with models using transmission simulations and flight data [13,14]. Generally, these approaches require location-specific inputs, limiting scalability and transferability. Alternatively, deaths seem to be a good indicator of true number of COVID-19 infections in the population [15]. Deaths have been used in past pandemics to estimate the true size of the pandemic given limited case identification [16]. The infection-fatality rate (IFR) of COVID-19 based on serological testing and comprehensive diagnostic testing has been shown to be between 0.7% and 1% [17–20], indicating that approximately 100 infections have occurred to each death. However, factors such as health system capacity, demography, and political regime impact the IFR [21]. Furthermore, heterogeneity in definitions of COVID-19-related deaths and testing strategies cause differences in completeness of the death count [22]. Thus, if deaths are used as an indicator of the true number of infections, adjustments may be necessary for testing, health system capacity, and government transparency.

A single, comprehensive metric that is scalable across all countries and territories would allow comparisons across states and identify ones most successful in more completely detecting the presence of infections in the population. It would also allow for statistical techniques, such as multiple linear regression, so researchers could comprehensively assess policy decisions in combination with other databases. Comprehensive metrics for COVID-19 and inter-country comparisons have been developed [23–25], but none that focus exclusively on testing or explicitly incorporate the true number of infections or detection rate, to the best of the authors’ knowledge. Thus, there is still a need for a single comprehensive metric that can overcome shortcomings in reported data to assess testing effectiveness during the COVID-19 pandemic.

Therefore, a single comprehensive metric that assesses testing effectiveness by incorporating an estimate of the true total number of COVID-19 infections in the population was developed using publicly reported data universally accessible across nearly all countries and territories. Model estimates of true period prevalence and detection rate were validated against comparable estimates in the literature. The metric was then used to assess factors associated with COVID-19 testing outcomes. We aimed to create a new tool for policymakers and researchers to comprehensively assess COVID-19 testing outcomes and identify effective policies.

## Methods and materials

### Data input

Data on COVID-19 were collected from the Worldometer website, which collects data directly from government communication channels and is managed by an international team of developers, researchers, and volunteers [26]. The data collected from Worldometer included total cases (C), total deaths (D), total recovered (R), active cases (A), population (P) (in millions), and total tests (T). These data have been reported explicitly or implicitly by the website since at least April 6, 2020. Prior to this date, subsets of these data were available.

Two other input data included the Global Health Security Index Detection and Reporting sub-index (I_sys_) [27] and the Economist Intelligence Unit Democracy Index (I_dem_) [28]. The I_sys_ assesses a health system’s capacity for early detection and reporting during epidemics of potential international concern. This index is available for 195 countries and has a scale from 0 to 100, where 100 indicates perfect detection and reporting. I_sys_ values were not available for Hong Kong and Taiwan. The value for Hong Kong, I_sys_=78, was imputed as the average between South Korea (Isys=92.1) and Singapore (Isys=64.5), which were assumed to have comparable health systems. Similarly, the value for Taiwan, I_sys_=81, was imputed as average between South Korea (I_sys_=92.1) and Japan (I_sys_=70.1). All other states without a value (n=17) were imputed as 42, which was the global average. I_dem_ is a projected measure of the degree of democracy; it is calculated for 167 countries and states and has a scale from 0 to 10, where 10 indicates the highest degree of democracy. This value was assumed to act as a proxy for transparency in data reporting. All states without a value (n=24) were imputed to be 5.4, the global average.

Countries and territories were included in the index if at least one case was reported, and the population was greater than or equal to 100,000. Data were accessed daily; however, this report presents the results for data accessed as of 00:00 GMT on June 3, 2020 (n=188).

### Definition of key indicators

Several key indicators were computed from the input data, representing important epidemiological indicators used in the analysis.

#### Case Fatality Rate (CFR)

The CFR is the proportion of total deaths, *D*, among closed cases (sum of *D* and *R*). However, some countries, including the Netherlands and United Kingdom, have not reported the number of recoveries, and others have not tracked recoveries in real-time. Additionally, the closed-case definition of CFR can overestimate the CFR in the early stages of an epidemic because of the relatively small number of closed cases [29]. Therefore, an alternative estimate of CFR was calculated as the ratio of deaths, *D*, to cases, *C*. Logically, the ratio of D:C is lower than D:(D+R) because C includes unresolved cases with unknown outcomes. Linear regression of these two ratios using data from the Worldometer [26] showed the relationship to be: D:(D+R) = 1.99*(D:C) (R^2^=0.58, n=178). Thus, the CFR used in our further analysis was the minimum of either the reported closed-case CFR or 2 times the ratio of *D*:*C* (Eq. 1).

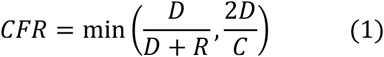

#### Test Positivity Rate (TPR)

TPR was computed as the ratio of cases, *C*, to tests, *T* (Eq. 2).

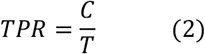

While the reported number of tests from specific countries or territories may represent multiple tests conducted on a single individual or even number of specimens, no adjustment was attempted to account for such heterogeneity in the various definitions of the TPR. In some cases, *T* was not available and thus TPR was not calculated.

#### Tests per Capita (TPC)

TPC was computed as the ratio of tests, *T*, to population (in millions), *P* (Eq. 3).

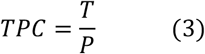

Similar to TPR, no adjustments were made to account for inter-country heterogeneity in definitions of *T*. In cases where *T* was not available, TPC was also not available.

### Estimating true number of infections and detection rate

It can be assumed that the reported number of cases, *C*, in a country represents a subset of the true number of infections. Some infections will go undetected, but as detection of cases increases, *C* will approach the true number of infections. Thus, the true number of infections (*Inf*) is some factor, *f*, higher than the cases that have been identified (Eq. 4).

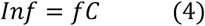

We conceptualized this factor, *f*, to be a function of the level of testing, the approach to testing (e.g., whether testing focuses on symptomatic, hospitalized, or general populations), and the quality and completeness of the data. We then used the CFR, TPR, TPC, I_sys_, and I_dem_ to formulate numerical values for *f*. Two separate formulations of *f* (*f*_*1*_ and *f*_*2*_) were defined, and the maximum of each of the two factors was used in the following analysis, as described below.

#### Factor 1 (f_1_)

Health system capacity and government transparency affect the completeness of data. To account for this, two multipliers were constructed: *m*_*sys*_ (Eq. 5) to adjust for the health system capacity using *I*_*sys*_—an indicator of a health system’s ability to detect and report cases; and *m*_*dem*_ (Eq. 6) to adjust for government transparency using *I*_*dem*_—an indicator of government transparency in reporting data.

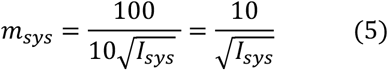

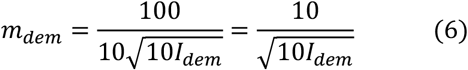

The mathematical relationships between the index and the multiplier in each of these equations follow declining relationships asymptotic to 1 and the principle that when health system capacity (I_sys_) or government transparency (I_dem_) are reduced, the multipliers increase, representing increased underreporting.

Additionally, another indicator of underreporting is a high ratio of deaths to cases, D:C. If all infections have been identified as cases and all cases resolve, then this ratio will approach the infection fatality rate (IFR), which has been estimated for COVID-19 to be around 1% or less [17]. That is, for every recorded death, at least 100 infections occurred. Thus, if the ratio of 100D:C exceeded 1, it was used together with m_sys_ and m_dem_ to determine *f*_*1*_, otherwise 1 was used and *f*_*1*_ was determined by the product of m_sys_ and m_dem_ only (Eq. 7).

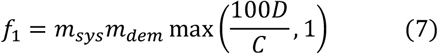

#### Factor 2 (f_2_)

Inadequate or low testing levels may also affect the completeness of the data. Therefore, a second factor incorporated data on testing (Eq. 8). This factor was comprised of two multipliers: *m*_*TPR*_ (Eq. 9) based on TPR—an indicator of adequate testing relative to disease prevalence—and *m*_*TPC*_ (Eq. 10) based on TPC—an indicator of adequate testing relative to the population.

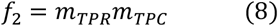

The World Health Organization (WHO) has suggested testing capacity is adequate when TPR is <10% [30]. If TPR is greater than 10%, it was inferred that increasingly more cases were undetected and that the multiplier m_TPR_ would be equal to the ratio of the TPR to that 10% benchmark (Eq. 9).

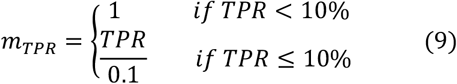

It was further assumed that if TPC was greater than 100,000 per million, or 10% of the entire population, TPC was extensive enough not to contribute to underreporting (i.e., *m*_*TPC*_ = 1).

Logically, as TPC decreases, the likelihood of undetected infections increases. The assumed relationship between the multiplier *m*_*TPC*_ and TPC followed a step-wise logarithmic relationship (Eq. 10).

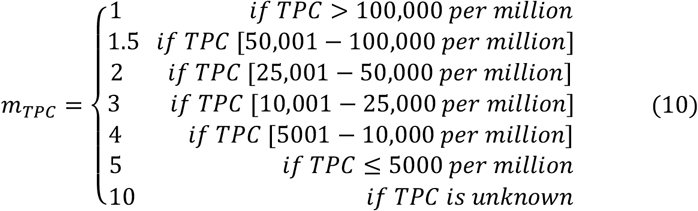

#### True number of infections

The true number of infections, *Inf*, was estimated as the product of *C* and the maximum of the two factors, whichever was highest (Eq. 11).

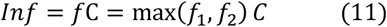

When possible, *Inf* was compared against national estimates from other empirical estimates in literature to validate the model.

#### Detection Rate (DR)

With an estimate of the true number of infections, it was possible to estimate what percentage of the infections (including asymptomatic) have been detected. The total number of cases, C, was divided by *Inf* to estimate the DR in each country or territory. The inverse of the factor, 1/*f*, represents the DR. Estimates of the DR were compared to DRs published in the literature.

### Calculation of the COVID-19 Testing Index

The COVID-19 Testing Index (CovTI) consists of four sub-indices, each based on a single key indicator. The key indicators used were the DR, TPR, CFR, and proportion of cases that are active (i.e., A/C). These indicators were chosen since they are commonly used as metrics and are computable with the methods described above. The relationship between each sub-index and its indicator was built on two principles. First, each sub-index was scaled from 0 to 100 with 0 representing the worst indicator value and 100 the best indicator value. Second, a square root mathematical relationship was used to scale the sub-index. A square root function was chosen as a simplistic way to reflect the complex reality, in which each marginal change of an indicator in the undesired direction (e.g., increase in TPR or decrease in DR) represents an increasingly higher risk of an uncontrolled COVID-19 epidemic or a worse testing response. These sub-indices were combined in a weighted average to compute CovTI.

#### Detection Rate sub-index (DR_si_)

If infections are undiagnosed, those individuals can actively spread the disease unknowingly. Without timely diagnosis, effective contact tracing cannot occur. Therefore, undiagnosed infections critically contribute to unchecked spread of COVID-19 and subsequently represent inadequate response. Thus, DR_si_ was given 40% weighting in computing the CovTI (Eq. 16). As the detection rate decreases, the DR_si_ decreases (Eq. 12).

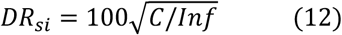

#### Test Positivity sub-index (TP_si_)

The TPR is a commonly used testing metric for COVID-19 [31]. A high TPR represents a reactive, rather than proactive, approach to testing. Accordingly, as TPR increases, the TP_si_ decreases (Eq. 13). If TPR was not available, a dummy value of 20 was used. TP_si_ was given 20% weighting in computing the CovTI.

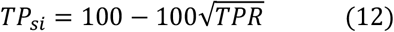

#### Case-Fatality sub-index (CF_si_)

The WHO has suggested that the CFR should be around 1 to 2 percent if the response is adequate. If CFR is higher, only severely infected patients are being diagnosed. Accordingly, if CFR increases beyond the minimum benchmark of 2%, CF_si_ decreases (Eq. 13). If the CFR is less than 2%, the CF_si_ is 100. CF_si_ was given 20% weighting. If the CFR was 0%, a dummy value of 50 was used.

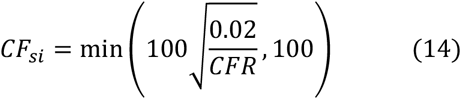

#### Active Case sub-index (ACsi)

Finally, a fourth sub-index accounts for how active the epidemic is in a country. If the epidemic is relatively active in the country, it is less likely the testing is adequate, and the increase reflects inadequate case identification. It provides a metric to incorporate progress as cases resolve. As the proportion of total cases that are active decreases, it reflects the epidemic is declining and passing, and the AC_si_ increases (Eq. 15). It was given a weight of 20%.

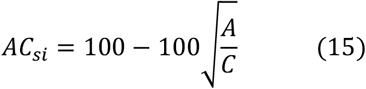

In the case of United Kingdom and the Netherlands that do not report *A*, the AC_si_ is not computable. In such cases, a dummy value of 50 was used.

#### COVID-19 Testing Index (CovTI)

The CovTI was calculated as the weighted average of the four sub-indices (Eq. 16) described above with a heavier weighting given to the DR_si_ due to the importance of undetected cases in driving uncontrolled epidemics and because it incorporates several factors into its derivation.

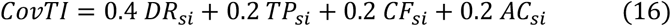

### Statistical analyses with CovTI

Five independent grouping variables were assessed for their effect on COVID-19 testing effectiveness by analyzing their association with CovTI (Table 1). Testing and contact tracing policy status were accessed from the Oxford COVID-19 Government Response Tracker [32] for May 13, 2020, which is three weeks prior to June 3, 2020, approximately the average time from symptom onset to death [19]. Islands were defined as any country that is an island, part of an island (co-island), or has limited land connections (limited land) or archipelago (see details in S1 Table). Crude bivariate analyses using two-tailed two-sample t-tests and one-way analysis of variance (ANOVA) were used to test whether the means between groups were different. A multiple linear regression (MLR) model was developed by using forced entry of all factors with p>0.20 in crude analysis. Factors were removed using backwards stepwise method (p >0.05) with Bayesian Information Criterion (BIC) used to assess model fit and overparameterization. Analyses were performed in Stata 14 [33].

**Table 1.** Definitions of grouping variables for multiple linear regression of COVID-19 Testing Index (n=163).

| Factor (Grouping Variables) | Operational Definition<sup>a</sup> | n (%) |
| --- | --- | --- |
| Testing Policy |  |  |
| No testing policy | OxCGRT Testing = 0 on May 13, 2020 | 6 (3.7) |
| Limited Testing | OxCGRT Testing = 1 on May 13, 2020 | 70 (42.9) |
| Symptomatic Testing | OxCGRT Testing = 2 on May 13, 2020 | 58 (35.6) |
| Open Public Testing | OxCGRT Testing = 3 on May 13, 2020 | 29 (17.8) |
| **Contact Tracing Policy** |  |  |
| No contact tracing | OxCGRT Contact Tracing = 0 on May 13, 2020 | 19 (11.7) |
| Limited contact tracing | OxCGRT Contact Tracing = 1 on May 13, 2020 | 64 (39.3) |
| Comprehensive contact tracing | OxCGRT Contact Tracing = 2 on May 13, 2020 | 80 (49.1) |
| **Geographical setting** |  |  |
| Island or island-like nation | Entirely on island(s) (including Australia) or parts of islands (e.g., Dominican Republic) or have very limited land connections (e.g., Hong Kong) | 28 (17.2) |
| Non-island nation | Not meeting the definition of island nation | 135 (82.8) |
| **Form of government** |  |  |
| Authoritarian government | $I_{\text{dem}} < 4$ | 48 (29.5) |
| Non-authoritarian federation | Constitutionally a federation with $I_{\text{dem}} \geq 4$ | 17 (10.4) |
| Non-authoritarian unitary state | Constitutionally not a federation with $I_{\text{dem}} \geq 4$ | 98 (60.1) |
| **Development status** |  |  |
| OECD<sup>b</sup> member | Member | 36 (22.1) |
| Non-OECD member | Not a member | 127 (77.9) |

1 <sup>a</sup>OxCGRT = Oxford COVID-19 Government Response Tracker [32]
<sup>b</sup>OECD = Organization for Economic Development

## Results

### True number of infections

Globally, the model estimated that approximately 68.2 million people have been infected in the period prior to June 3, 2020, compared to the reported 6.47 million cases (mean multiplier factor, *f*, =10.5, range = 1.5 to 165). In other words, for every reported case it is estimated that 9.5 infections have gone unreported.

### Detection rate

Globally, the DR was estimated to be 9.5% (range = 0.6-66.6%). The countries estimated to have the highest detection rates were Australia, Singapore, Iceland, and Hong Kong (Table 2), each estimated to have detected at least 50% of all infections.

**Table 2.** Top 10 countries and territories according to highest implied detection rate (data 00:00 GMT June 3, 2020).

| Global Rank | Country or Territory | Multiplier Factor, $f$ | Implied Detection Rate |
| --- | --- | --- | --- |
| 1 | Australia | 1.50 | 66.6% |
| 2 | Singapore | 1.61 | 62.3% |
| 3 | Iceland | 1.68 | 59.7% |
| 4 | Hong Kong | 2.00 | 50.0% |
| 5 | Kuwait | 2.31 | 43.2% |
| 6 | New Zealand | 2.51 | 39.9% |
| 7 | Israel | 2.61 | 38.3% |
| 8 | Latvia | 2.62 | 38.1% |
| 9 | Malta | 2.84 | 35.2% |
| 10 | Bahrain | 2.93 | 34.2% |

### Comparison to previous estimates

This model’s estimates were compared against historical estimates in the literature (Table 3). The results showed that this model’s estimates were similar to previous estimates at comparable time periods (R^2^=0.97). In many cases the estimates of true number of cases and DR closely matched previous estimates, and in most cases the estimates were within the 95% confidence interval of previous estimates.

**Table 3.** Comparison of total cases and detection rate estimates with other similar estimates in literature [34] [35].

|  |  | Other Estimates[34][35] |  | This Study's Estimate |  |
| --- | --- | --- | --- | --- | --- |
| Date | Country | Estimated Period Prevalence [95% CI] | Implied Detection Rate [95% CI] | Estimated Period Prevalence | Implied Detection Rate |
|  |  | % of total population | % of infections | % of total population | % of infections |
| 14 April 2020 [34] | Australia | 0.04 [0.03-0.09] | 59 [28-90] | 0.07 | 33 |
|  | Canada | 1.0 [0.67-2.2] | 6.7 [3.1-10] | 0.25 | 28 |
|  | South Korea | 0.06 [0.04-0.13] | 35 [16-53] | 0.06 | 33 |
|  | USA | 2.6 [1.7-5.6] | 6.8 [3.2-10] | 1.5 | 13 |
| 4 May<br>2020<br>[35] | Austria | 0.76 [0.59-0.98] | 23.4 [18-30] | 0.85 | 20 |
|  | Belgium | 8.0 [6.1-11] | 5.4 [3.9-7.1] | 9.7 | 4.4 |
|  | Denmark | 1.0 [0.81-1.4] | 17 [12-21] | 0.92 | 17.6 |
|  | France | 3.4 [2.7-4.3] | 7.6 [6.0-9.6] | 4.9 | 5.3 |
|  | Germany | 0.85 [0.66-1.1] | 23 [18-30] | 0.95 | 21 |
|  | Italy | 4.6 [3.6-5.8] | 7.5 [6.0-9.6] | 6.2 | 5.6 |
|  | Norway | 0.46 [0.34-0.61] | 32 [24-44] | 0.51 | 28 |
|  | Spain | 5.5 [4.4-7.0] | 9.6 [7.5-12] | 6.5 | 8.1 |
|  | Sweden | 3.7 [2.8-5.1] | 6.0 [4.4-8.0] | 2.9 | 7.4 |
|  | Switzerland | 1.9 [1.5-2.4] | 18 [14-23] | 2.8 | 12.4 |
|  | UK | 5.1 [4.0-6.5] | 5.3 [4.2-6.8] | 4.8 | 5.6 |

### COVID-19 Testing Index

#### Comparison between Countries

Countries in the top quartile of CovTI had lower TPR, lower CFR, lower proportion of active cases, and higher DR (Fig 1). The top 10 countries according to the index were dominated by island nations and states that are effectively islands (e.g., Hong Kong) (Table 4a). Among non-island nations, Thailand, Slovakia, and Israel had the highest CovTI (Table 4b). Full results are reported in Supporting Information (S1 and S2 Tables).

**Table 4.** COVID-19 Testing Index (CovTI) and sub-indices a) among all countries and territories assessed (n=188) and b) among only non-island nations (n=143). DR_si_, TP_si_, CF_si_, AC_si_, as described in text with percentages indicating degree of weighting. Data per 00:00 GMT June 3, 2020. Complete data set in S1 Table.

| **Global Rank** | **Country or Territory** | **CovTI** | **DR<sub>si</sub>**<br>**(40%)** | **TP<sub>si</sub>**<br>**(20%)** | **CF<sub>si</sub>**<br>**(20%)** | **AC<sub>si</sub>**<br>**(20%)** |
| --- | --- | --- | --- | --- | --- | --- |
| 1 | Iceland | 86.8 | 77.3 | 82.8 | 100.0 | 96.7 |
| 2 | Australia | 86.0 | 81.6 | 93.0 | 100.0 | 73.9 |
| 3 | New Zealand | 83.3 | 63.1 | 92.7 | 100.0 | 97.4 |
| 4 | Hong Kong | 82.5 | 70.7 | 92.7 | 100.0 | 78.2 |
| 5 | Brunei | 76.7 | 50.3 | 91.5 | 100.0 | 91.6 |
| 6 | Malta | 75.8 | 59.4 | 90.6 | 100.0 | 69.7 |
| 7 | Thailand | 75.5 | 50.0 | 91.4 | 100.0 | 86.2 |
| 8 | Slovakia | 74.8 | 55.9 | 90.8 | 100.0 | 71.7 |
| 9 | Israel | 74.4 | 61.8 | 82.8 | 100.0 | 65.5 |
| 10 | Taiwan | 73.5 | 44.7 | 92.2 | 100.0 | 85.7 |

**b)**
| **Global Rank** | **Country or Territory** | **CovTI** | **DR<sub>si</sub>**<br>**(40%)** | **TP<sub>si</sub>**<br>**(20%)** | **CF<sub>si</sub>**<br>**(20%)** | **AC<sub>si</sub>**<br>**(20%)** |
| --- | --- | --- | --- | --- | --- | --- |
| 7 | Thailand | 75.5 | 50.0 | 91.4 | 100.0 | 86.2 |
| 8 | Slovakia | 74.8 | 55.9 | 90.8 | 100.0 | 71.7 |
| 9 | Israel | 74.4 | 61.8 | 82.8 | 100.0 | 65.5 |
| 11 | South Korea | 73.3 | 57.7 | 88.9 | 88.8 | 73.3 |
| 14 | Malaysia | 72.6 | 57.7 | 88.1 | 100.0 | 59.5 |
| 16 | Georgia | 72.1 | 57.7 | 88.4 | 99.8 | 56.7 |
| 17 | Jordan | 71.6 | 57.7 | 93.8 | 100.0 | 48.9 |
| 18 | Montenegro | 71.3 | 44.9 | 82.1 | 84.9 | 100.0 |
| 21 | Palestine | 69.8 | 50.0 | 90.0 | 100.0 | 58.9 |
| 22 | Kyrgyz Republic | 69.1 | 57.7 | 87.3 | 100.0 | 42.5 |

**Fig 1.**
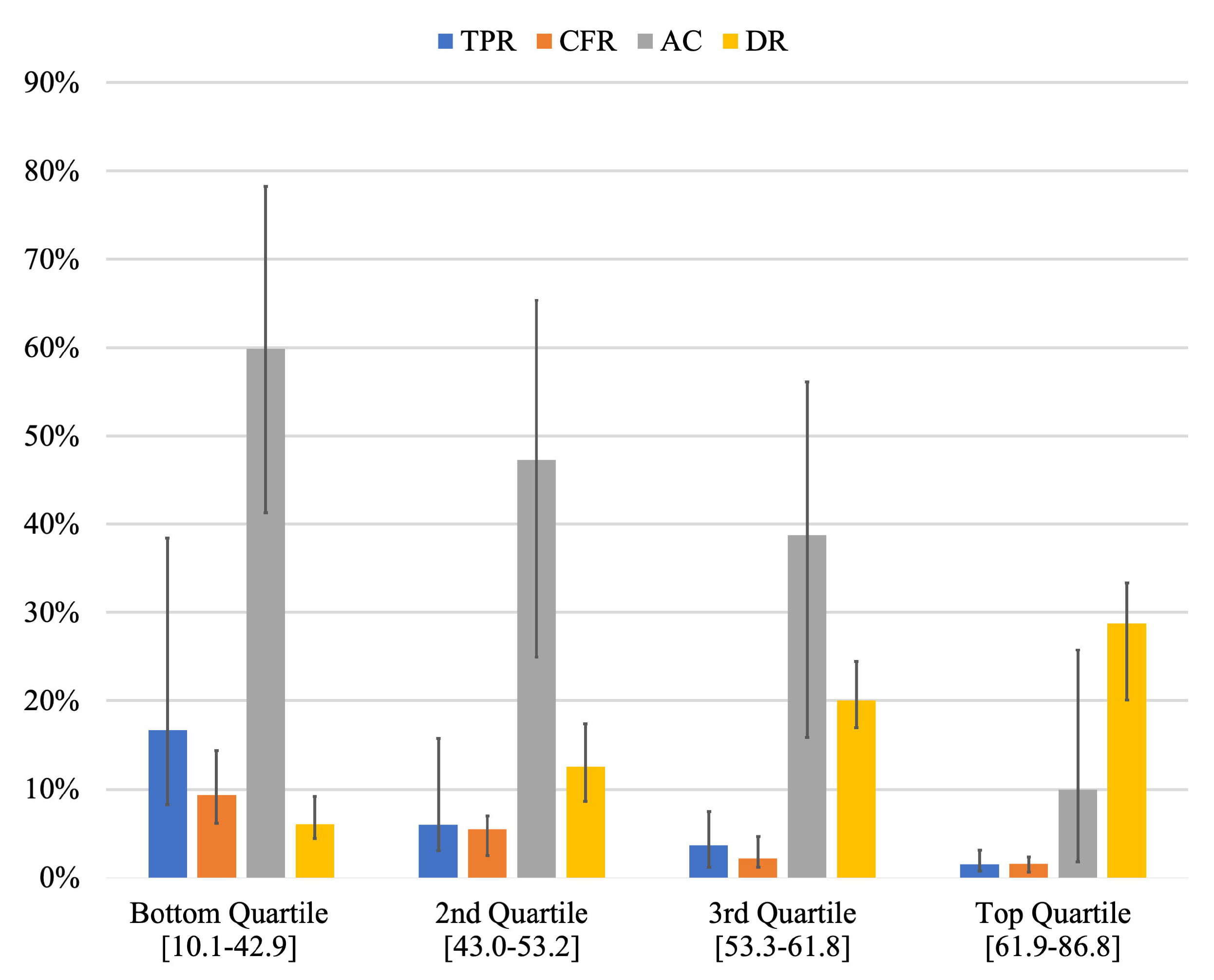
Comparison of the medians of test positivity rate (TPR), case fatality rate (CFR), proportion of active cases (AC), and detection rate (DR) among the quartiles of COVID-19 Testing Index (CovTI). Bottom quartile includes the lowest 25% of CovTI values (n=47), 2^nd^ quartile includes CovTI values between 25^th^ and 50^th^ percentiles (n=47), 3^rd^ quartile includes CovTI values between 50^th^ and 75^th^ percentiles (n=47), and the top quartile includes highest CovTI values above 75^th^ percentile (n=47). Values in brackets indicate the range of CovTI values in each quartile. Error bars represent interquartile range. Data per 00:00 GMT June 3, 2020.

#### Temporal Comparison

Comparing the index from April 20 to June 3, the global average of CovTI increased nearly 10 points from 43.2 to 52.2 (Fig 2). The index in most countries (89.9%) increased over this time period. Some countries, such as Australia, increased substantially, while other countries, such as Germany and USA, increased comparably to the global average increase. Other countries (10.1%) had index values that decreased, including Russia, Canada, and Brazil.

**Fig 2.**
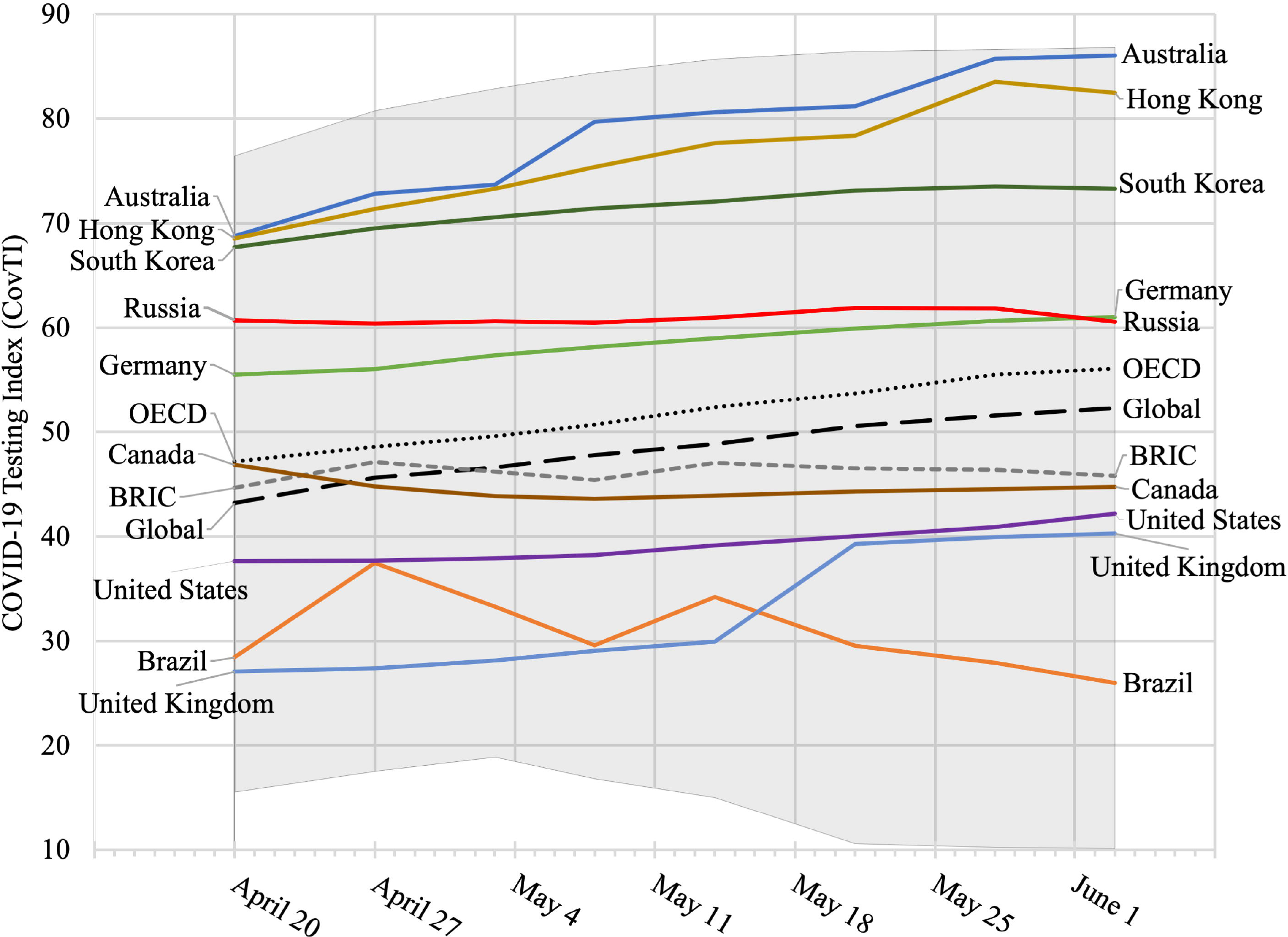
Trend of COVID-19 Testing Index among selected countries and groups. OECD = Organization for Economic Cooperation and Development. BRIC = Brazil, Russia, India, and China. Shaded grey area represents full range of CovTI values.

#### Variable Analysis

Bivariate analyses showed that testing policy and contact tracing policy were significantly associated (p<0.0001) with CovTI (Table 5). Additionally, the trend demonstrated that increasing levels of testing and contact tracing was associated with improved CovTI. Island nations were significantly more effective than non-islands, and significant differences in CovTI were found between forms of government. OECD members had better COVID-19 testing outcomes, but the differences were not significant (p=0.11).

**Table 5.** **Comparison of differences in COVID-19 Testing Index between countries and states with different testing and tracing policies, geographical settings, forms of government and economic development status (n=163)**.

| Variable | n (%) | CovTI<br>$\bar{x}$ (95% CI) | Test<br>statistic<sup>a</sup> | p-value |
| --- | --- | --- | --- | --- |
| **Testing policy** |  |  | 10.4 | <0.0001 |
| No testing policy | 6 (3.7) | 30.2 (18.1-42.3) |  |  |
| Limited testing | 70 (42.9) | 49.5 (46.2-52.8) |  |  |
| Symptomatic testing | 58 (35.6) | 54.7 (51.1-58.2) |  |  |
| Open public testing | 29 (17.8) | 59.7 (55.6-63.8) |  |  |
| **Contact tracing policy** |  |  | 14.5 | <0.0001 |
| No contact tracing | 19 (11.7) | 40.8 (33.8-47.9) |  |  |
| Limited contact tracing | 64 (39.3) | 49.7 (46.3-53.1) |  |  |
| Extensive contact tracing | 80 (49.1) | 57.4 (54.6-60.1) |  |  |
| **Geographical setting** |  |  | 3.82 | 0.0002 |
| Island or island-like | 28 (17.2) | 61.4 (55.6-67.2) |  |  |
| Non-island | 135 (82.8) | 50.6 (48.3-52.9) |  |  |
| **Form of government** |  |  | 3.05 | 0.05 |
| Non-authoritarian unitary state | 98 (60.1) | 54.5 (51.8-57.3) |  |  |
| Authoritarian government | 48 (29.4) | 50.1 (45.8-54.4) |  |  |
| Non-authoritarian federation | 17 (10.4) | 47.0 (40.4-53.7) |  |  |
| **Economic development status** |  |  | 1.60 | 0.11 |
| OECD Member | 36 (22.1) | 55.8 (50.8-60.7) |  |  |
| Non-OECD Member | 127 (77.9) | 51.5 (49.1-54.0) |  |  |

<sup>a</sup> t-statistic from two-way two-sample t-test with equal variance or F-statistic from one-way

All factors were entered into the initial MLR model. The final model included all factors except economic development. The MLR showed that testing policy had the largest effect on testing outcomes, whereby widespread open testing was associated with a 23.8-point increase in CovTI compared to no testing policy (Table 6). Contact tracing, centralized governments, and islands were also associated with improved CovTI values.

**Table 6.**
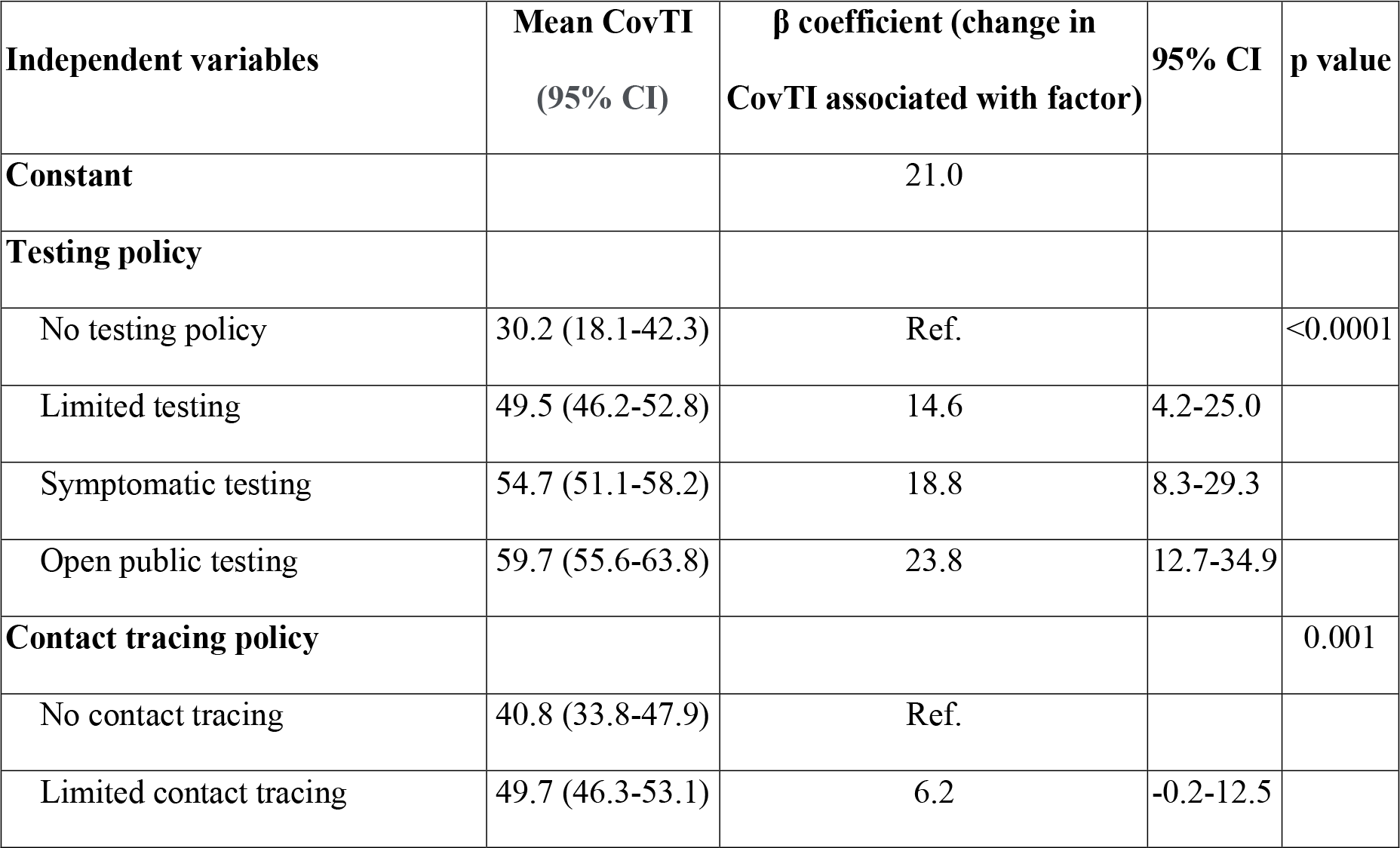
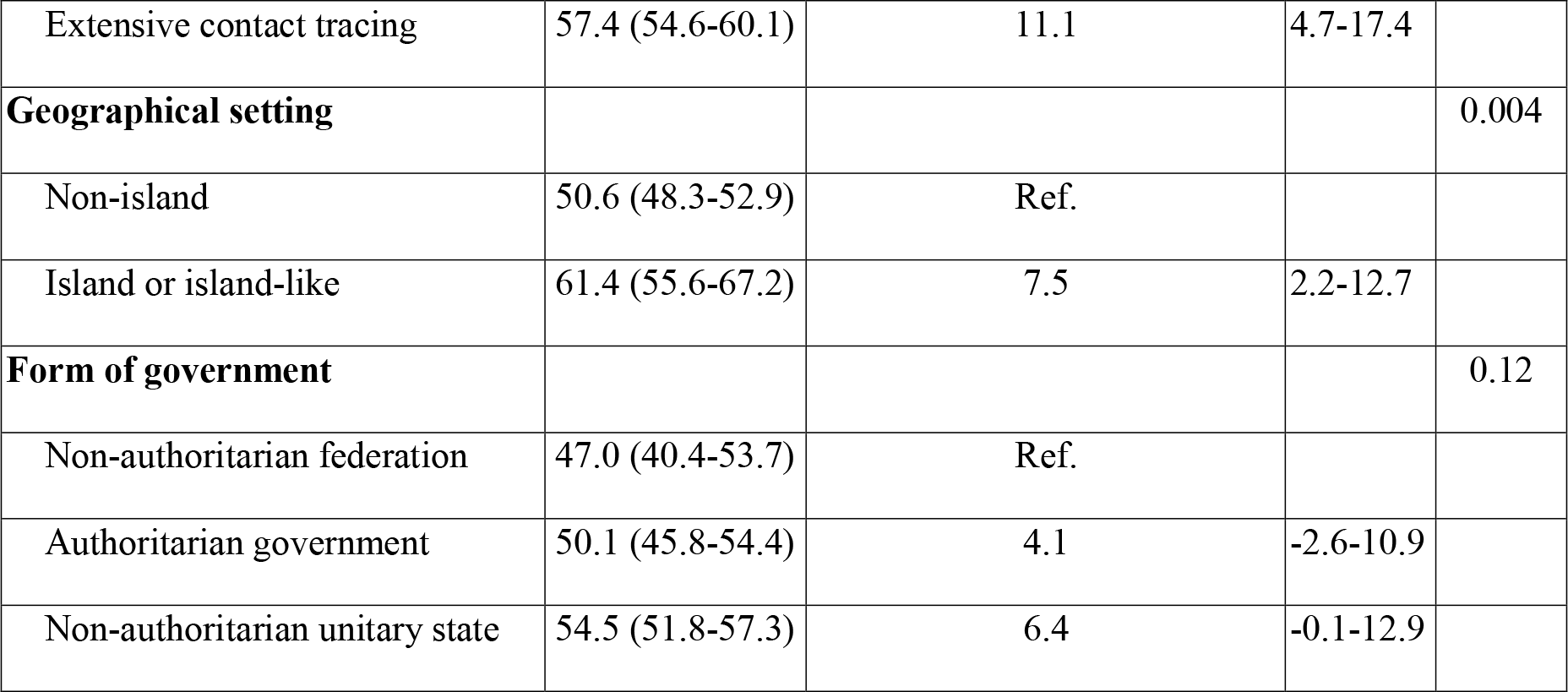
**Multiple linear regression analysis F(8, 154) = 9**.**06 (p<0**.**0001), adjusted R**^**2**^ **= 0**.**28 of factors associated with COVID-19 Testing Index from final model (n=163) (Data from 00:00 GMT June 3, 2020)**.

| Independent variables | Mean CovTI<br>(95% CI) | $\beta$ coefficient (change in<br>CovTI associated with factor) | 95% CI | p value |
| --- | --- | --- | --- | --- |
| **Constant** |  | 21.0 |  |  |
| **Testing policy** |  |  |  |  |
| No testing policy | 30.2 (18.1-42.3) | Ref. |  | <0.0001 |
| Limited testing | 49.5 (46.2-52.8) | 14.6 | 4.2-25.0 |  |
| Symptomatic testing | 54.7 (51.1-58.2) | 18.8 | 8.3-29.3 |  |
| Open public testing | 59.7 (55.6-63.8) | 23.8 | 12.7-34.9 |  |
| **Contact tracing policy** |  |  |  | 0.001 |
| No contact tracing | 40.8 (33.8-47.9) | Ref. |  |  |
| Limited contact tracing | 49.7 (46.3-53.1) | 6.2 | -0.2-12.5 |  |
| Extensive contact tracing | 57.4 (54.6-60.1) | 11.1 | 4.7-17.4 |  |
| **Geographical setting** |  |  |  | 0.004 |
| Non-island | 50.6 (48.3-52.9) | Ref. |  |  |
| Island or island-like | 61.4 (55.6-67.2) | 7.5 | 2.2-12.7 |  |
| **Form of government** |  |  |  | 0.12 |
| Non-authoritarian federation | 47.0 (40.4-53.7) | Ref. |  |  |
| Authoritarian government | 50.1 (45.8-54.4) | 4.1 | -2.6-10.9 |  |
| Non-authoritarian unitary state | 54.5 (51.8-57.3) | 6.4 | -0.1-12.9 |  |

## Discussion

We developed a novel comprehensive metric, entitled the CovTI, that attempts to measure effectiveness of testing during the COVID-19 pandemic at the country level and is derived from key epidemiological indicators computable from data available across nearly all countries, as reported on the Worldometer website and other similar databases [26]. Previous research has assessed government response [23] and suggested specific indicators to facilitate inter-country comparisons [24]. However, this is the first published metric to comprehensively assess testing outcomes with a focus on detection/underreporting. The results showed that testing and contact tracing independently contribute to better detection and should be used in conjunction with one another. The results showed that the parsimonious model yielded estimates of true period prevalence consistent with previous estimates as shown below.

### National testing policy impact on testing outcomes

Testing effectiveness, as indicated by CovTI, was strongly associated with contact tracing and testing policies. This validates the model’s ability to track COVID-19 testing outcomes and further provides confirmation that countries that prioritize policies and dedicate resources specifically to testing and contact tracing have effectively reduced underreporting and improved testing-related health outcome metrics, such as the CFR.

The results also show that increasingly more inclusive policies for testing (for example, open public testing vs. symptomatic testing) and contact tracing (e.g., extensive vs. limited) yielded progressively better detection rates and testing outcomes. Therefore, countries should aim to increasingly expand these efforts. Additionally, contact tracing and testing independently contributed to better outcomes, even after adjusting for other factors. Hence, policies should ensure increased testing is paired with increased contact tracing capacity.

High values in the proposed index showed correlation with countries that have been recognized for their success in responding to the COVID-19 pandemic, including Taiwan, Australia, Iceland, and South Korea. These countries are recognized for their high testing rates and comprehensive contact tracing programs [36,37]. Thus, this analysis provided quantitative evidence supporting policy recommendations to facilitate strong national leadership, expand diagnostic capacity, rapidly enact comprehensive contact tracing, and proactively test for COVID-19 [36,37].

### Extrinsic factors affecting testing outcomes

This analysis found that island nations had better testing effectiveness outcomes compared to non-island nations. Geographical isolation is an obvious advantage to controlling infectious disease [38]. The results also showed an association of higher CovTI with authoritarian and unitary forms of government. This result suggests an association between better testing outcomes and more centralized forms of government. Previous research has shown associations between the level of democracy and CFR, where the time trajectories of CFR increase more steeply for democracies than for autocracies, i.e. democracies have higher CFR than autocracies [21]. A higher CFR indicates poorer testing effectiveness, i.e. lower CovTI (Fig 1). Here we show that centralized governments have higher CovTI and lower CFR, which is consistent with Sorci et al. [21]. Thus, extrinsic factors affect a country’s ability to implement testing strategies. Combining this metric with other databases that can account for other possible factors, such as trust in government institutions, demographics, or urban/rural distribution, could further elucidate other extrinsic factors related to COVID-19 testing.

### Estimate of true number of cases and detection rate

A parsimonious empirical model was used to estimate the true number of infections and inferred detection rate by back-calculating from reported deaths with adjustments for health system capacity, government transparency, and testing levels. The results were comparable to more complex modeling approaches based on similar principles [34,35]. The findings suggested that nearly 90% of global infections remain unreported, which is consistent with previous estimates showing that true number of infections are many times higher than reported cases [9–11]. In addition, other authors have used different models to estimate the true number of cases as a percentage of the population in various European countries. For example, period prevalence in Italy was estimated at 4 percent in April [39] and 4.4 percent in France in May [40]. Several of the hardest affected countries early in the pandemic had an estimated period prevalence between 3 and 7.5 percent [41]. These estimates were generally consistent with our estimates (Table 3).

Seroprevalence of antibodies is often touted as a reliable means to estimate period prevalence and past exposure to SARS-CoV-2. Efforts are underway in June 2020 across England, Germany, and the United States, among others, to randomly sample the population and determine country-level seroprevalence. However, due to the specificity of the serological tests, false positives can substantially affect the accuracy of the results[42] Therefore, models and parsimonious estimates may continue to play an important role in estimating the true number of infections.

### Herd immunity

The estimates of this model further agree with models in the hardest hit countries, such as Italy or Spain, which estimate that more than five percent of the population in those countries have been infected [41]. However, such a low proportion of the population presumably with antibodies is far from conferring herd immunity that may inhibit future disease transmission. It is important to note that, while these proportions are much higher than the officially reported cases, they do not represent herd immunity—a concept considered important to fully reopening society. Although herd immunity depends on the basic reproductive number (R_0_) [43], which varies with effectiveness of interventions, some estimates specify a threshold of 50 to 60 percent seroprevalence to achieve herd immunity [44], while others, accounting for differential susceptibility, estimate the threshold may be as low as 20 percent [45]. Nevertheless, these estimates suggest that herd immunity is not yet occurring at the national level of the countries analyzed.

### Excess deaths

Reported deaths are an important indicator. However, excess deaths (i.e., total mortality in excess of seasonal averages) are substantially higher than the reported COVID-19 deaths in many locations, including Brazil, Jakarta, New York City, and Ecuador [46]. These excess deaths suggest such locations have not accurately captured deaths related to COVID-19 in official figures [47]. Different definitions of attributable deaths substantially affect data. For example, Russia had previously used a very limited definition for inclusion determined via autopsy that does not count many deaths even if the patient previously tested positive for SARS-CoV-2 [48]. On the other hand, some countries have opted to exhaustively include any presumptive death to COVID-19 in official data. Belgium has included unconfirmed deaths within their COVID-19 death total [49]. In such a case, COVID-19 deaths are greater than excess deaths [46]. These examples of limited or conservative death definitions impact this model’s estimates. Nevertheless, substantial differences between excess deaths and deaths attributed to COVID-19 may indicate insufficient testing or policy decisions that systematially exclude some deaths. While proxy factors, including the adjustments for government transparency and health system capacity, were included to mitigate this factor, it is likely the model estimates will not sufficiently adjust in cases of excessive deaths across such wide ranging scenarios. Including data on excessive deaths could improve the validity of the model; however, such data is usually reported with a lag of several weeks or not reported at all.

### Sources of uncertainty

Several assumptions were made to generate this model. Most notably, the model assumes specific and universal relationships between deaths and total number of infections, implying an inherent IFR. Advances in therapeutics and differences in health system capacity will influence this rate, though [21]. In addition, several factors including age, sex, hypertension, diabetes, and blood groups are known to affect the mortality and hospitalization rates [50–56]. Future analyses may combine CovTI with databases that include these factors. The model also assumes specific relationships between proxy indicators, such as the Global Health Security Index and Democracy Index, and data outcomes. While the direction of the relationship is arguably evident, the magnitude and shape of the relationship is unknown. Furthermore, this model aims to be parsimonious (i.e., not introducing excessive parameters or uncertainty) and is, by nature, deterministic. The decision to include a minimum number of variables and data was strategic, but a stochastic approach could better illustrate the uncertainty and sensitivity to the above assumptions. The model also reported estimates of total number of cases and detection rates. These values should be used cautiously as a comparative tool, rather than exact values.

## Conclusion

This report described a novel comprehensive metric (COVID-19 Testing Index, CovTI) that evaluates the overall effectiveness of COVID-19 testing in the current pandemic using real-time publicly reported data among 188 countries and territories. The metric incorporated case-fatality rate, test positivity rate, proportion of active cases, and an estimate of detection rate based upon reported death data by adjusting for heterogeneity in testing levels, health system capacity, and government transparency. The estimated detection rate of COVID-19 aligned satisfactorily with previous empirical and epidemiological models. National policies that allow open public testing and extensive contact tracing were significantly associated with higher values of CovTI, which reflects improvements in the estimated detection rate. Extrinsic factors, including geographic isolation and centralized forms of government, were also shown to be associated with higher COVID-19 testing outcomes. Countries should commit to expanding policies on testing and contact tracing in order to reduce levels of undetected infections and reduce disease transmission. Applications of this metric include combining it with different databases to identify other factors that affect testing outcomes or using it to temporally track a holistic measure of testing outcomes at the national level.

## Data Availability

Our data was obtained from publicly available data, including:
Worldometer COVID-19 (April 20, 2020 and June 3, 2020) (https://www.worldometers.info/coronavirus/) provided data on Total Cases, Total Deaths, Total Recovered cases, Active Cases, Total Tests and population.
The Economist Intelligence Unit Democracy Index (2019) (https://www.eiu.com/topic/democracy-index) provided data for the Democracy Index.
Global Health Security Index (October 2019) (https://www.ghsindex.org/report-model/) provided data for the Detection and Reporting Country Score.
Oxford Coronavirus Government Response Tracker (May 13, 2020) (https://ourworldindata.org/grapher/covid-19-testing-policy, https://ourworldindata.org/grapher/covid-contact-tracing) provided data on COVID-19 Testing Policies, which countries do COVID-19 contact tracing?
A complete table of our June 3, 2020 dataset is reported in Supplementary Material S1 Table.
The full dataset that was analyzed and reported in Supplementary Material S2 Table and is available by request. A complete dataset, including historical data that was not analyzed in this paper, can also be made available.

https://www.eiu.com/topic/democracy-index

https://www.ghsindex.org/report-model/

https://ourworldindata.org/grapher/covid-19-testing-policy

https://ourworldindata.org/grapher/covid-contact-tracing

## Supporting information

**S1 Table. Raw data inputs and computed values for the COVID-19 Testing Index (CovTI) on June 3, 2020**. Raw data inputs, key epidemiological indicators, multipliers, factors, and sub-indices used to compute CovTI on June 3, 2020 among eligible countries and territories (n=188). Further details of each variable are described in the text. C=cases, D=total deaths, R=total recovered, A=active cases, P= population (in millions), T=total tests, CFR=case fatality rate, TPR=test positivity rate, TPC=tests per capita, m_sys_=health system capacity multiplier, m_dem_= democracy multiplier, m_TPR_= test positivity rate multiplier, m_TPC_= per capita testing multiplier. f1=factor 1, f2=factor 2, Inf= true number of infections, Prev=Estimated Period Prevalence = Inf /P, Act=proportion active cases=A/C, DR=detection rate, DRsi= Detection Rate sub-index, TPsi=Test Positivity sub-index, CFsi= Case-Fatality sub-index ACsi= Active Case sub-index, CovTI=COVID-19 Testing Index. OECD= Organization for Economic Development member, BRIC= Brazil, Russia, India, and China.

**S2 Table. Dataset for bivariate and multiple linear regression analyses**. CovTI as of June 3, 2020; island status; form of government; OECD membership; COVID-19 testing policy as of May 13, 2020; and COVID-19 contact tracing policy as of May 13, 2020 among eligible countries and territories with complete data (n=163). Further details of each variable are described in the text.

